# Clinical characteristics and early outcomes in patients with COVID-19 treated with tocilizumab at a United States academic center

**DOI:** 10.1101/2020.05.13.20100404

**Authors:** Casey A. Rimland, Camille E. Morgan, Griffin J. Bell, Min K. Kim, Tanner Hedrick, Ashley Marx, Brian Bramson, Heidi Swygard, Sonia Napravnik, John L. Schmitz, Shannon S. Carson, William A. Fischer, Joseph J. Eron, Cynthia L. Gay, Jonathan B. Parr

## Abstract

We describe early outcomes in 11 COVID-19 patients treated with the IL-6 receptor inhibitor tocilizumab. While C-reactive protein decreased, neither clinical improvement nor reduced temperature or oxygen requirements was observed in most patients. Our findings contrast with prior reports and raise questions about tocilizumab use in severe COVID-19.

## INTRODUCTION

A hyperinflammatory state is a posited driver of severe COVID-19 [1]. Elevations of IL-6 have been described in acute respiratory distress syndrome and cytokine release syndrome (CRS) [2,3]. Recently, IL-6-driven inflammation in severe COVID-19 was suggested, as increased levels were associated with mortality [4]. IL-6 blockade via tocilizumab, a humanized monoclonal antibody that targets the IL-6 receptor, has been proposed as a treatment for COVID-19 [5].

Peer-reviewed reports of tocilizumab use for COVID-19 are largely limited to retrospective case series from China [6,7] and Italy [8,9], as well as case reports [10–13]. Xu *et al*. reported improved inflammatory markers, oxygen requirements, and fever in 21 patients after tocilizumab treatment [6]. Luo *et al.’s* description of 15 patients included more critically ill patients and more measured results, with 20% mortality [7]. A prospective, open-label study from Italy reported clinical improvement and increased survival after tocilizumab [8], while a second study in Italy found no significant clinical improvement or mortality benefit following tocilizumab [9]. Given the paucity of data on tocilizumab use in COVID-19 in the United States, we evaluated 11 patients treated with tocilizumab for COVID-19. We detail their clinical characteristics before and after treatment and reconsider tocilizumab use in severe COVID-19.

## METHODS

We performed a retrospective cohort study of patients with COVID-19 treated with tocilizumab at UNC Medical Center between March 21st and April 25th, 2020. Clinical data was extracted from the medical record. COVID-19 was diagnosed using nasopharyngeal swabs and reverse-transcriptase PCR (RT-PCR). IL-6 testing was conducted at Mayo Clinic Laboratories (Rochester, MN) using an electrochemiluminescence assay. Baseline IL-6 values for two patients were from a multi-cytokine panel (lower-limit of detection [LOD] of 5.0 pg/mL); all others were from a standalone IL-6 assay (LOD 1.8 pg/ml). Disease severity was defined according to the World Health Organization (WHO) COVID-19 clinical status scale with minor modification [14]. Descriptive statistics were calculated using JMP 15 and Wilcoxon signed rank test performed in GraphPad Prism 8. Data was visualized using the *tidyverse* package in R 3.6.0. The study was approved by the UNC Institutional Review Board.

## RESULTS

### Patient population

Eleven patients with confirmed SARS-CoV-2 infection treated with tocilizumab for COVID-19 were identified and included. Demographic, clinical, and laboratory data are displayed in **Table 1**. Six (55%) patients had an unknown exposure history. Median time from symptom onset to admission was 8 days (IQR=5-11). Most patients were male (82%, n=9); non-white (73%, n=8); non-substance users (82%) with a median age of 59 years old (IQR=48-65). Patients had a median of 2 (IQR=1-6) comorbidities, including hypertension (73%, n=8), type 2 diabetes mellitus (36%, n=4), and lung disease (27%, n=3). Almost all were overweight (36%, n=4) or obese (55%, n=6). Five (46%) took an angiotensin converting enzyme inhibitor (ACEI) or angiotensin receptor blocker (ARB) at baseline. Seven (64%) were transferred from outside hospitals, all within one day of initial presentation. Five (46%) were diagnosed with COVID-19 by SARS-CoV-2 RT-PCR as outpatients prior to presentation.

**Table 1.**
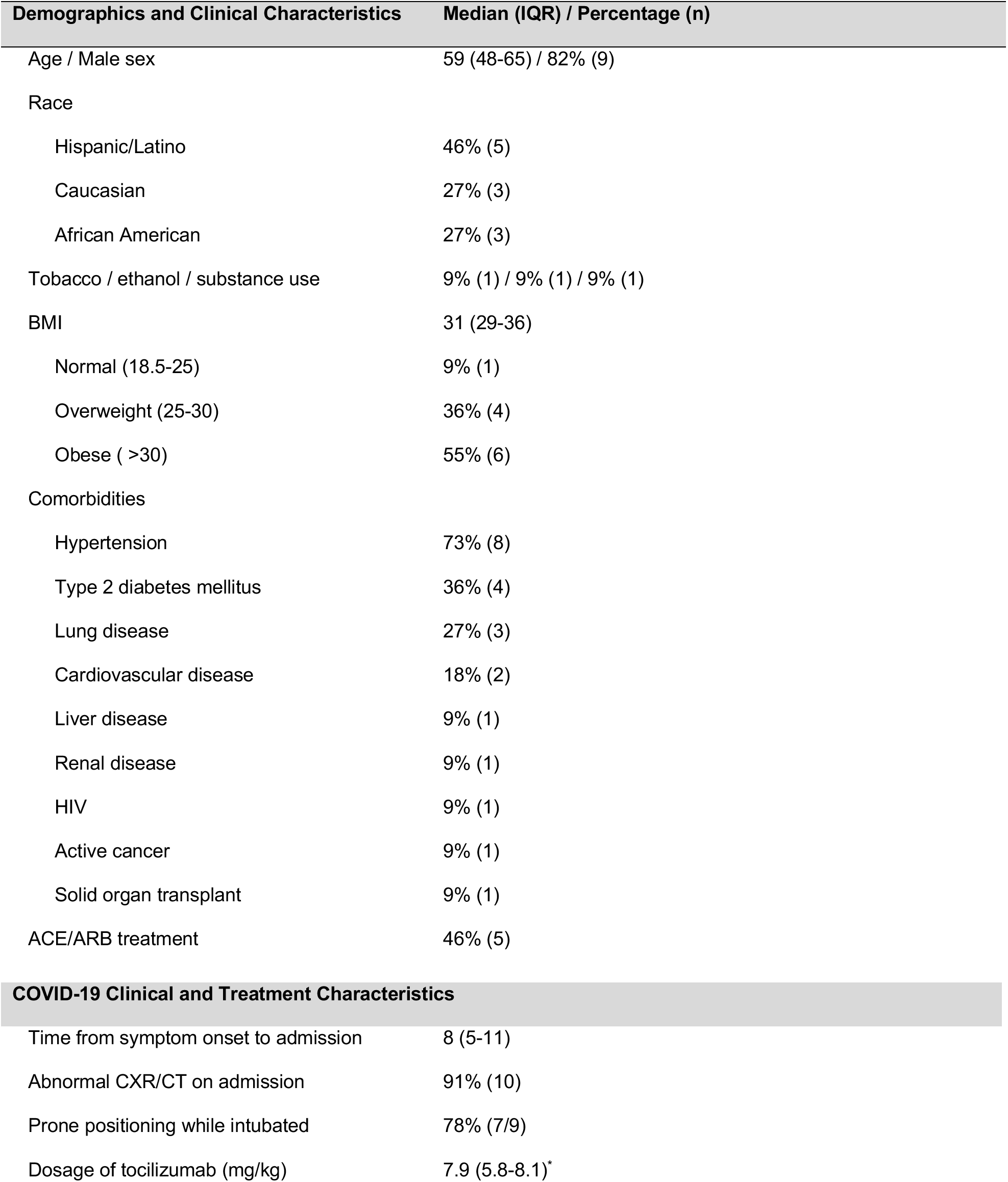
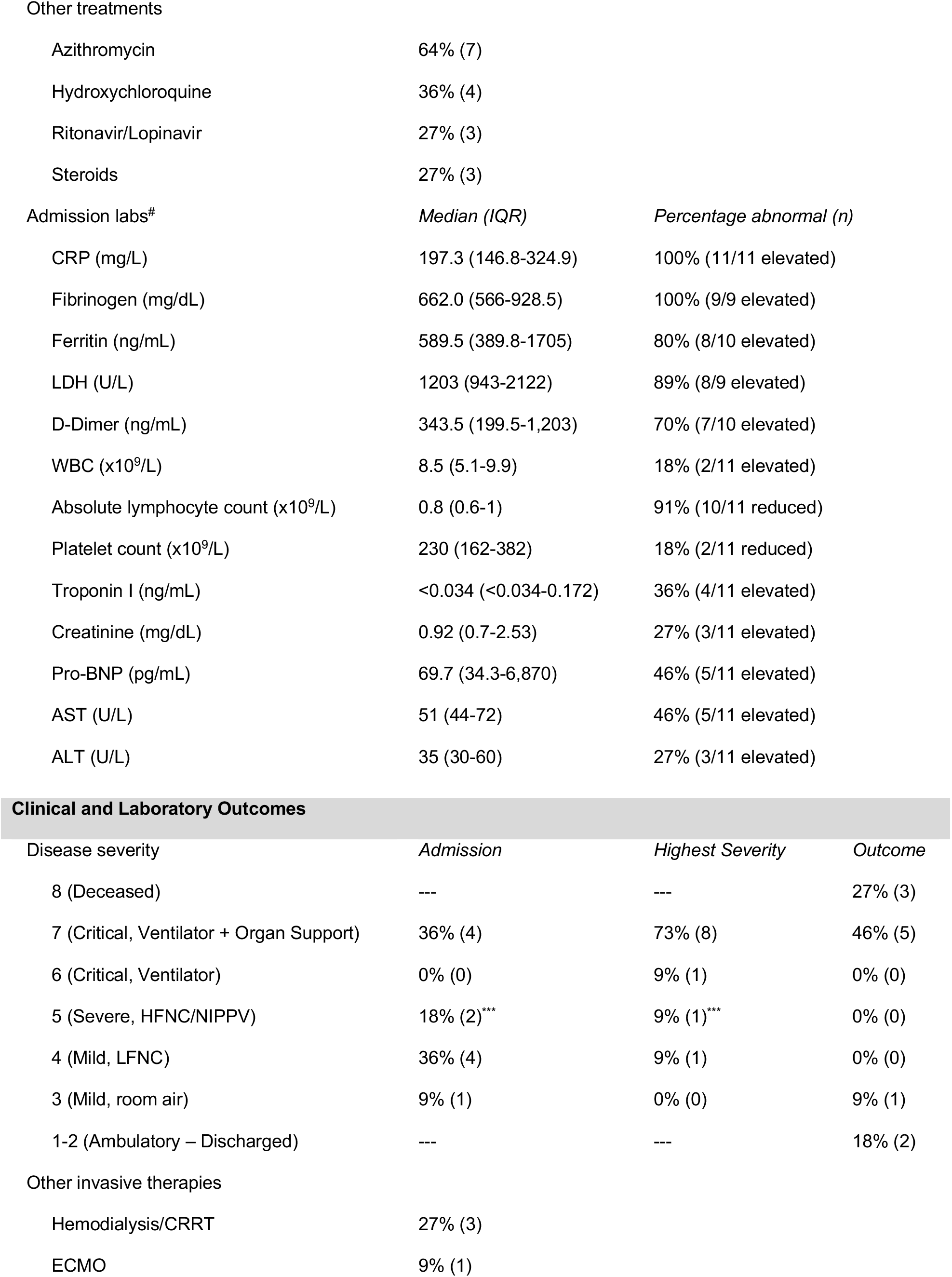
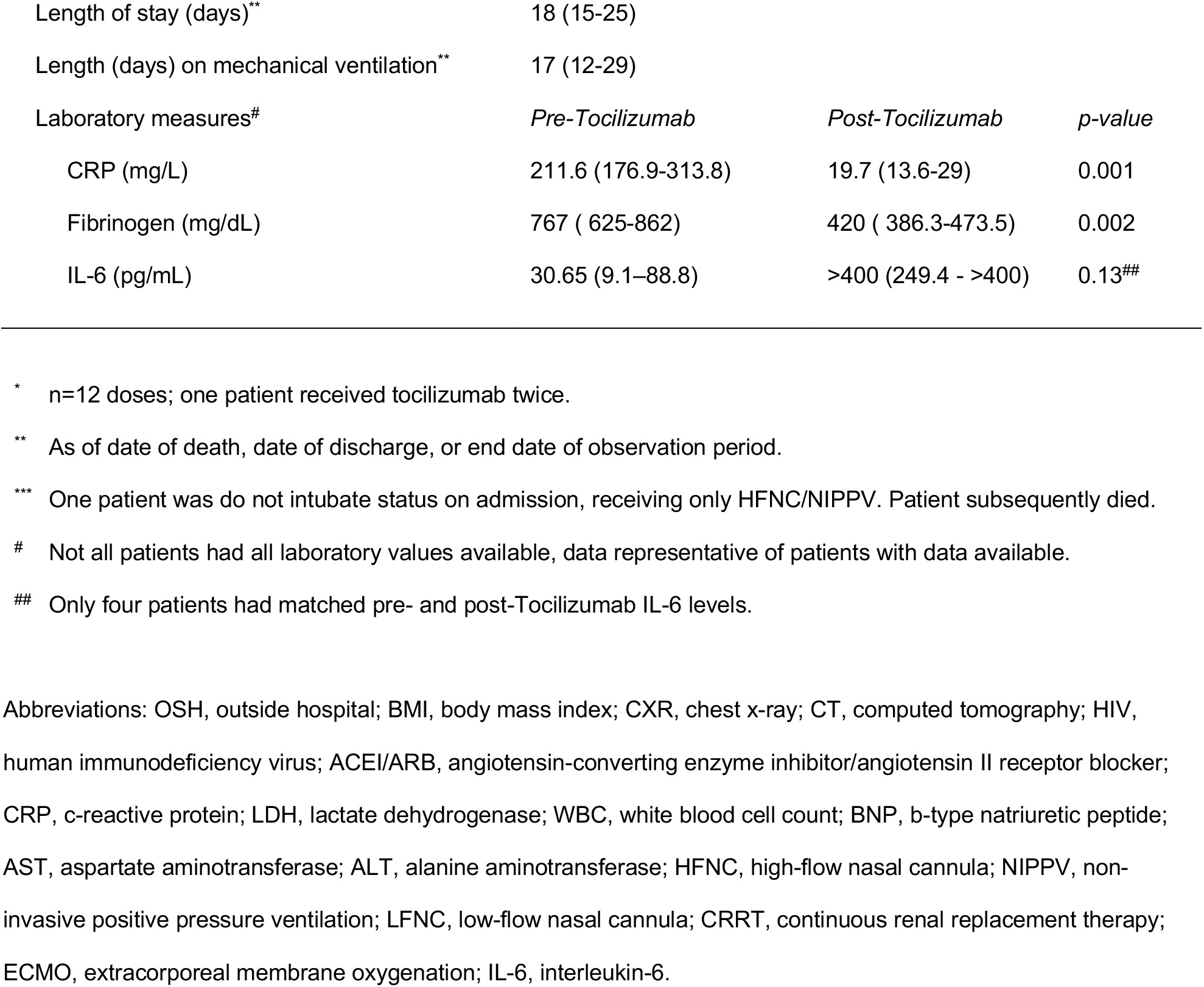
Patients with COVID19 treated with tocilizumab demographics and clinical characteristics (n=11).

| Demographics and Clinical Characteristics | Median (IQR) / Percentage (n) |
| --- | --- |
| Age / Male sex | 59 (48-65) / 82% (9) |
| Race |  |
| Hispanic/Latino | 46% (5) |
| Caucasian | 27% (3) |
| African American | 27% (3) |
| Tobacco / ethanol / substance use | 9% (1) / 9% (1) / 9% (1) |
| BMI | 31 (29-36) |
| Normal (18.5-25) | 9% (1) |
| Overweight (25-30) | 36% (4) |
| Obese ( >30) | 55% (6) |
| Comorbidities |  |
| Hypertension | 73% (8) |
| Type 2 diabetes mellitus | 36% (4) |
| Lung disease | 27% (3) |
| Cardiovascular disease | 18% (2) |
| Liver disease | 9% (1) |
| Renal disease | 9% (1) |
| HIV | 9% (1) |
| Active cancer | 9% (1) |
| Solid organ transplant | 9% (1) |
| ACE/ARB treatment | 46% (5) |
| **COVID-19 Clinical and Treatment Characteristics** |  |
| Time from symptom onset to admission | 8 (5-11) |
| Abnormal CXR/CT on admission | 91% (10) |
| Prone positioning while intubated | 78% (7/9) |
| Dosage of tocilizumab (mg/kg) | 7.9 (5.8-8.1)* |
| Other treatments |  |
| Azithromycin | 64% (7) |
| Hydroxychloroquine | 36% (4) |
| Ritonavir/Lopinavir | 27% (3) |
| Steroids | 27% (3) |

| Admission labs<sup>#</sup> | Median (IQR) | Percentage abnormal (n) |
| --- | --- | --- |
| CRP (mg/L) | 197.3 (146.8-324.9) | 100% (11/11 elevated) |
| Fibrinogen (mg/dL) | 662.0 (566-928.5) | 100% (9/9 elevated) |
| Ferritin (ng/mL) | 589.5 (389.8-1705) | 80% (8/10 elevated) |
| LDH (U/L) | 1203 (943-2122) | 89% (8/9 elevated) |
| D-Dimer (ng/mL) | 343.5 (199.5-1,203) | 70% (7/10 elevated) |
| WBC (x10<sup>9</sup>/L) | 8.5 (5.1-9.9) | 18% (2/11 elevated) |
| Absolute lymphocyte count (x10<sup>9</sup>/L) | 0.8 (0.6-1) | 91% (10/11 reduced) |
| Platelet count (x10<sup>9</sup>/L) | 230 (162-382) | 18% (2/11 reduced) |
| Troponin I (ng/mL) | <0.034 (<0.034-0.172) | 36% (4/11 elevated) |
| Creatinine (mg/dL) | 0.92 (0.7-2.53) | 27% (3/11 elevated) |
| Pro-BNP (pg/mL) | 69.7 (34.3-6,870) | 46% (5/11 elevated) |
| AST (U/L) | 51 (44-72) | 46% (5/11 elevated) |
| ALT (U/L) | 35 (30-60) | 27% (3/11 elevated) |

Clinical and Laboratory Outcomes
| Disease severity | Admission | Highest Severity | Outcome |
| --- | --- | --- | --- |
| 8 (Deceased) | --- | --- | 27% (3) |
| 7 (Critical, Ventilator + Organ Support) | 36% (4) | 73% (8) | 46% (5) |
| 6 (Critical, Ventilator) | 0% (0) | 9% (1) | 0% (0) |
| 5 (Severe, HFNC/NIPPV) | 18% (2)<sup>***</sup> | 9% (1)<sup>***</sup> | 0% (0) |
| 4 (Mild, LFNC) | 36% (4) | 9% (1) | 0% (0) |
| 3 (Mild, room air) | 9% (1) | 0% (0) | 9% (1) |
| 1-2 (Ambulatory – Discharged) | --- | --- | 18% (2) |
| Other invasive therapies |  |  |  |
| Hemodialysis/CRRT | 27% (3) |  |  |
| ECMO | 9% (1) |  |  |
| Length of stay (days)** | 18 (15-25) |  |  |
| Length (days) on mechanical ventilation** | 17 (12-29) |  |  |
| Laboratory measures# | *Pre-Tocilizumab* | *Post-Tocilizumab* | *p-value* |
| CRP (mg/L) | 211.6 (176.9-313.8) | 19.7 (13.6-29) | 0.001 |
| Fibrinogen (mg/dL) | 767 ( 625-862) | 420 ( 386.3-473.5) | 0.002 |
| IL-6 (pg/mL) | 30.65 (9.1–88.8) | >400 (249.4 - >400) | 0.13## |

\* n=12 doses; one patient received tocilizumab twice.
\*\* As of date of death, date of discharge, or end date of observation period.
\*\*\* One patient was do not intubate status on admission, receiving only HFNC/NIPPV. Patient subsequently died.

### Baseline laboratory and imaging characteristics

On admission, most patients had significantly elevated inflammatory markers including elevated CRP (100%, n=11/11), fibrinogen (100%, n=9/9), ferritin (80%, n=8/10), LDH (88.9%, n=8/9), and D-dimer (70%, n=7/10). Most patients had normal total white blood cell counts (WBC) (81.82%, n=9/11); two patients had elevated total WBC (18.2%). However, almost all patients presented with low absolute lymphocyte counts (90.9%, n=10/11). Several patients presented with laboratory signs of additional organ damage. Four patients had abnormal troponin I levels (36.4%), five patients had elevated pro-BNP (45.5%), three patients had elevated creatinine (27.3%), and two patients had both elevated AST and ALT (18.2%). Almost all patients had abnormal chest imaging on admission (91%, n=10).

### Hospital course

Most patients had severe or critical COVID-19 by modified WHO criteria (55%, n=6) at the time of admission (**Table 1**). All patients required supplemental oxygen on admission, including five (46%) via low-flow nasal cannula (LFNC, ≤ 10L/min), two (18%) via high-flow nasal cannula (HFNC), and four (36%) via mechanical ventilation. During the observation period, eight (73%) were critically ill requiring mechanical ventilation and vasopressors, one (9%) was critically ill requiring mechanical ventilation alone, one (9%) with do-not-intubate status received HFNC only, and one (9%) had mild disease requiring LFNC. Ten of the 11 (91%) received at least one dose of an anti-microbial or immunomodulating treatment during their hospitalization.

### Outcomes after tocilizumab

Tocilizumab was administered a median of one day (IQR 1-4) from admission and nine days (IQR 7-14) after symptom onset. Ten patients received a single tocilizumab dose, and one received two doses. Median dosage was 7.9 mg/kg. Two of six patients with available baseline IL-6 levels had low levels, one below the assay’s LOD (≤5 pg/mL) and another of 10.4 pg/mL. Four of five with available IL-6 levels after tocilizumab had IL-6 levels above the upper limit of quantification. IL-6 increased after tocilizumab in all four patients with matched baseline and post-tocilizumab testing.

CRP and fibrinogen decreased significantly after tocilizumab (**Figure 1A**, median CRP 211.6 vs. 19.7 mg/L [*p*=0.001] and fibrinogen 767 vs. 420 mg/dL [*p*=0.002] pre- and 5 days post-tocilizumab, respectively). Other laboratory markers did not show clear trends (**Figure 1B**). Temperature did not show obvious improvement (**Figure 1C**), and only three (27%) showed improvement in oxygen requirements (**Figure 1D**). At end of follow-up, patients were observed for a median of 17 days (range=7-34) after initial tocilizumab dose. Three died (27%); five (45%) remained in the intensive care unit (ICU) on mechanical ventilation, receiving vasopressors or hemodialysis; one (9%) was transferred from the ICU to floor status and weaned to room air; and two (18%) were discharged home without oxygen. Seven patients (64%) had minimally elevated liver function tests after tocilizumab administration. Two were diagnosed with ileus and two with bacterial pneumonia after tocilizumab administration; no serious adverse events were observed.

**Figure 1.**
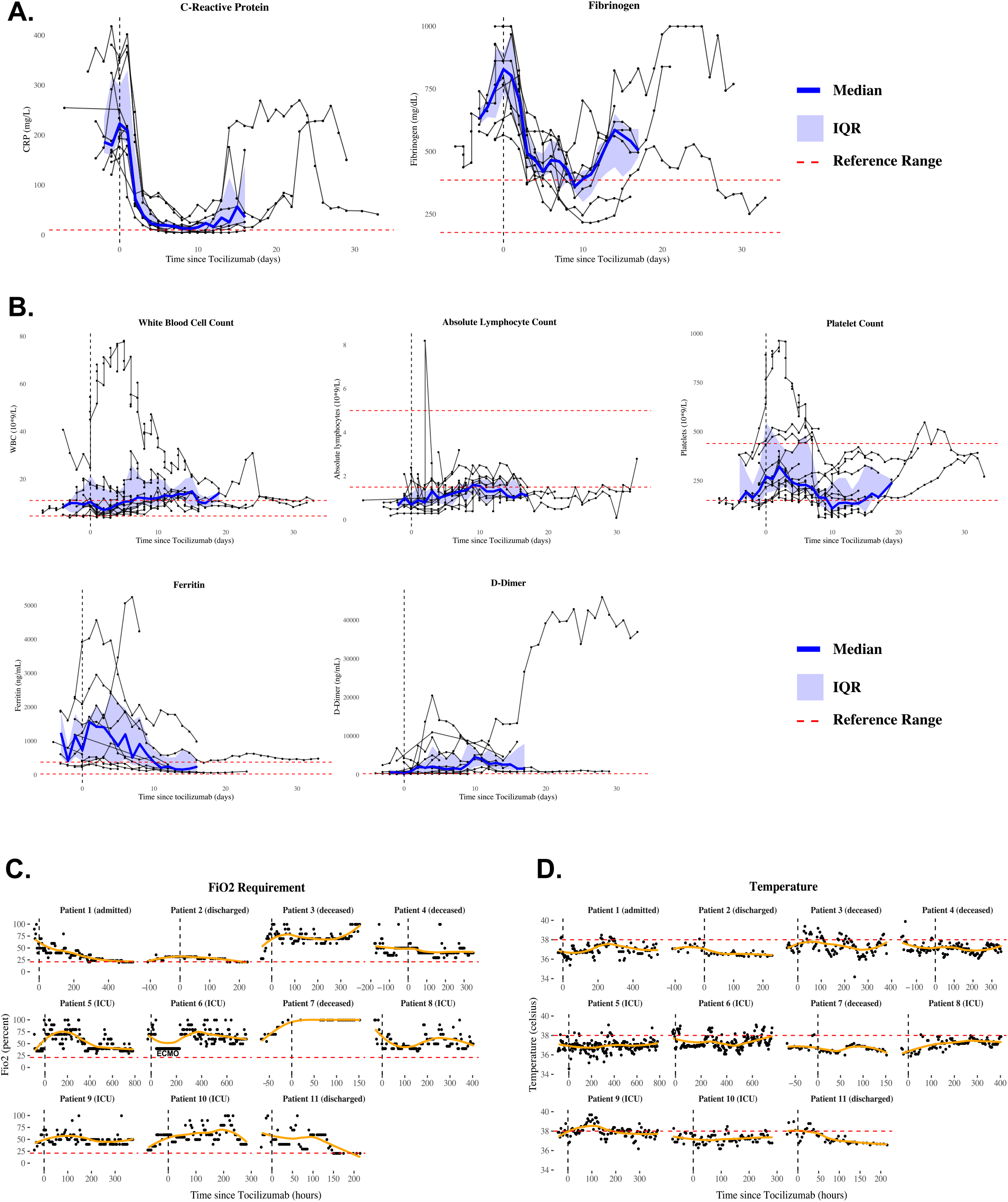
Laboratory and clinical outcomes of patients treated with tocilizumab. (**A-B**) Laboratory trends following tocilizumab. Medians (blue lines), interquartile ranges (IQR, blue shading), and reference ranges (dashed red lines). (**C-D**) Oxygen requirements and temperature trends, displayed using locally estimated scatterplot smoothing (orange lines). Patient #6 was treated with two doses of tocilizumab; time 0 refers to the time of initial administration.

## DISCUSSION

While tocilizumab was recently described as a promising therapy for severe COVID-19, our findings raise questions about its use in critically ill patients [7]. We report data from patients with COVID-19 treated with tocilizumab at a United States academic center, almost all of whom required ICU care. CRP and fibrinogen improved after tocilizumab administration as previously reported [6–8], but improvement in disease severity or mortality were not observed. These findings, in addition to low baseline IL-6 levels observed in some patients, raise questions about tocilizumab use in patients presenting with severe or critical COVID-19, especially without baseline IL-6 data.

Important differences exist between our cohort and those from China and Italy [6–9]. Three of four studies had few patients requiring mechanical ventilation (n=2/21, n=4/32, and n=5/63 respectively) [6,8,9]; the fourth study did not report specific oxygen requirements but did include more patients reported as critically ill (n=7/15) [7]. Our cohort included higher rates of hypertension (73% versus 40% in Luo *et al*., 37% in Campochiaro *et al*., and 38% in Sciascia *et al.);* a majority were overweight or obese (91%). We also observed lower baseline IL-6 levels compared to those described in these cohorts, although IL-6 assays were conducted in different laboratories. While Sciascia *et al*. attribute observed survival benefit of tocilizumab treatment to early administration in their Italian cohort [8], we did not observe the same clinical improvement, despite tocilizumab administration within a median of one day of hospitalization. Our findings also demonstrate the challenge of defining CRS in COVID-19, especially in the absence of standardized IL-6 assays and limited data to support IL-6 thresholds for tocilizumab use.

Our findings contrast with most previous reports and do not provide clear evidence of clinical improvement after tocilizumab treatment in patients with severe COVID-19. Several explanations can be considered. First, our patients were more severely ill than previous cohorts. It is possible tocilizumab was administered too late to offer benefit. Second, our patient cohort was more racially and ethnically diverse and had more comorbidities, notably obesity and hypertension, than those in China and Italy. Third, improvements observed in prior, less severely ill cohorts, might have occurred as part of the normal disease process, with or without tocilizumab.

In conclusion, tocilizumab reduces inflammatory markers (CRP, fibrinogen), but it is unclear whether its activity leads to equally dramatic effects on clinical outcomes. Randomized controlled trials are essential to determine when, or if, tocilizumab should be used in severe COVID-19.

## Data Availability

Data available on request.

## ACKNOWLEDGMENTS

The authors would like to thank the nurses; laboratory staff; and allied health providers involved in the care of these patients. We also thank Carlton Moore and Hannah Little for assistance with initial exploration of data.

